# Association of Obesity with Kidney Function Outcomes in Heart Failure with Preserved Ejection Fraction

**DOI:** 10.1101/2025.05.19.25327963

**Authors:** Lixia Deng, Hocine Tighiouart, Tatsufumi Oka, Marcelle L. Tuttle, Brian C. Downey, Ethan J. Rowin, Jennifer E. Ho, Mark Sarnak, Wendy McCallum

## Abstract

**Background:** Kidney dysfunction is highly prevalent in heart failure with preserved ejection fraction (HFpEF). It poses therapeutic challenges and is associated with worse clinical outcomes. Obesity is increasingly recognized as a key factor in HFpEF pathogenesis, yet its impact on kidney function remains unclear.

**Methods:** We conducted a retrospective analysis using data from the Treatment of Preserved Cardiac Function Heart Failure with an Aldosterone Antagonist (TOPCAT) trial. Obesity was examined using body mass index (BMI), waist circumference (WC) and waist to height ratio (WHtR). The primary outcome was a decline in estimated glomerular filtration rate (eGFR) by >30% from baseline. Single eGFR decline was defined as meeting this threshold with one qualifying follow-up eGFR; persistent eGFR decline was defined by meeting it on two consecutive qualifying eGFR values. Univariable and multivariable Cox proportional hazards regression models were performed, modeling each exposure as a continuous variable and as quartiles.

**Results:** A total of 1765 patients were included (mean age 72±10 years, median eGFR of 60.8 [IQR 47.8, 76.1] ml/min/1.73m^2^) with a median follow up of 3.3 years. There were 690 (39.1%) patients who met the definition of single eGFR>30% decline, and 459 (26%) patients who met the definition of persistent eGFR>30% decline. Compared to the lowest quartile, there was a significantly higher risk of >30% eGFR decline in the highest quartile of BMI (HR=1.26 [95% CI 1.00, 1.60]), WC (HR=1.35 [1.06, 1.71]) and WHtR (HR=1.27 [1.00, 1.61]), with similar trends in continuous analyses. All associations were attenuated and no longer met statistical significance when using the outcome of persistent kidney function decline.

**Conclusions:** Obesity is independently associated with declines in kidney function in patients with HFpEF. Associations were similar but attenuated and no longer met statistical significance for persistent declines in kidney function, suggesting that perhaps obesity is a risk factor for fluctuations in eGFR in HFpEF.

## Introduction

It is widely accepted that the presence of kidney dysfunction among patients with heart failure with preserved ejection fraction (HFpEF) is associated with worse clinical outcomes and higher risk of mortality. The coexistence of HFpEF and chronic kidney disease (CKD) presents additional diagnostic and therapeutic challenges due to diminished capacity for sodium excretion leading to complications such as diuretic resistance and volume overload. There can also be limitations of available therapeutic treatments due to low kidney function, contributing to adverse clinical outcomes.^1^ Despite its clinical significance, the pathophysiology and risk factors driving declines in kidney function among patients with HFpEF remain incompletely understood.

Studies have shown that obesity in HFpEF can be associated with higher cardiac filling pressures, acceleration of inflammatory pathways and increased renin angiotensin aldosterone system (RAAS) activity.^2^ In other words, prior studies have hinted at numerous pathways outside of traditional risk factors—hypertension and diabetes—that could link obesity in HFpEF with increased risk of declines in kidney function.

Upward of 80% of patients with HFpEF have obesity or are overweight, yet whether obesity is associated with decreased kidney function in this subtype of heart failure is not fully understood.^3^ With the paradigm shift in approach toward HFpEF, obesity is increasingly recognized as playing a key role in the pathogenesis through systemic proinflammatory state and coronary endothelial dysfunction. ^4^ In this study, we sought to investigate whether obesity is a potential risk factor driving worsening kidney function in HFpEF.

## Method

### Study Population

We utilized data from the Treatment of Preserved Cardiac Function Heart Failure with an Aldosterone Antagonist (TOPCAT) trial obtained from the National Heart, Lung, and Blood Institute Biologic Specimen and Data Repository Information Coordinating Center. The TOPCAT trial was a multicenter, randomized double-blind trial investigating the effect of spironolactone versus placebo in patients with HFpEF, conducted from 2006 to 2012. The trial included adults ≥ 50 years of age meeting at least one sign and one symptom of heart failure, left ventricular ejection fraction (LVEF) ≥45%, with history of heart failure hospitalization within the last 12 months, or with N-Terminal Pro-B-Type Natriuretic Peptide (NT proBNP) ≥360 pg/mL or b-type natriuretic peptide (BNP) ≥100 pg/mL. Patients were randomized to receive spironolactone or placebo. The primary outcomes of the trial were a composite outcome of cardiovascular death, aborted cardiac arrest or hospitalization for management of heart failure.^5^ For the current analysis, data was limited to that from the Americas due to previously reported regional heterogeneity in data from Eastern Europe. Patients with a baseline creatinine and at least one additional measurement obtained during follow-up were included. The current analysis was deemed exempt from review for the Tufts Medical Center Institutional Review Board.

### Exposure

The exposures of interest included obesity and abdominal adiposity characterized in three different ways: 1) elevated body mass index (BMI), 2) elevated waist circumference (WC), and 3) elevated waist to height ratio (WHtR). For BMI, the World Health Organization’s threshold of BMI ≥30 kg/m^2^ was used. High waist circumference (WC) is defined by ≥102 cm in men and ≥88 cm in women as previously defined.^6^ High WHtR is defined by ≥0.5 in both men and women.^7^ Each measure of obesity was examined as a continuous variable and as quartiles.

### Outcome

The outcomes of interest included decline of eGFR by >30% from baseline eGFR, which is a widely accepted surrogate kidney endpoint in the CKD literature.^8^ We defined this endpoint in two ways: 1) based on a single eGFR data point, where patients meet the endpoint with one measure of eGFR decline by >30%; and 2) based on two eGFR data points, where patients with two consecutive measures meeting the endpoint were defined as persistent eGFR decline. The exception was if the kidney endpoint was reached on the last measure of eGFR available, a confirmatory measure was not required. The eGFR was estimated using creatinine and the CKD-EPI 2021 formula.^9^ The eGFR was measured at the time of randomization, then every four months. Only measurements of kidney function according to pre-specified protocol were included.

### Covariates

We included covariates based on prior literature including sex, race, age, prior history of hypertension, diabetes mellitus, baseline eGFR, use of angiotensin-converting enzyme inhibitors (ACEI), angiotensin receptor blocker (ARB), use of a non-potassium sparing diuretic, and randomization arm to spironolactone vs. placebo.

### Statistical Analyses

Results are presented as mean ± SD for normally distributed values, or median (IQR) for non-normal distribution. Baseline characteristics were compared between those meeting definitions of obesity and those without obesity by all three metrics using student *t* test for continuous variables, and chi-square test for categorical variables. Kaplan Meier estimates of the probabilities of each kidney outcome were performed separately according to those with obesity and without obesity by each metric (BMI, WC, WHtR). The association between BMI, WC and WHtR as continuous variables and quartiles with kidney endpoints was examined using multivariable Cox proportional hazard regression model incorporating the aforementioned covariates. The time at risk began at the time of randomization into the trial. Patients were censored at the time of their last available eGFR. Death was treated as a censoring event.

Two subgroups were also further investigated. As women tend to have a higher prevalence of both obesity and HFpEF compared to men, and given previously observed sex differences in obesity-related traits, we evaluated associations between obesity and kidney outcomes within subgroups of female vs male patients. Due to the known acute eGFR lowering effect of spironolactone, we also explored subgroups by spironolactone vs placebo as well as effect modification by randomization arm.^10^ Several sensitivity analyses were performed. First, all models were repeated substituting the >30% eGFR decline threshold with >40% eGFR decline. We also performed competing risk analyses using Fine and Gray models treating death as a competing risk for eGFR decline. All analyses were performed using SPSS Statistics (Version 28.0.0.0, IBM Corp, Armonk, New York) and R language (Version 3.3.1, R Foundation for Statistical Computing, Vienna, Austria)

## Results

### Baseline characteristics

A total of 1765 patients from TOPCAT with both a randomization and at least one follow-up eGFR were identified and included in the analysis. Baseline characteristics by obesity defined by BMI are shown in Table 1, and by obesity defined by WC and WHtR in Supplemental Tables S1 and S2. There were 631 (35.8%) patients in the normal weight group (BMI<30 kg/m^2^) and 1135 (64.2%) in the obese group (BMI≥30 kg/m^2^). Patients in the obese group were younger (69±15 vs 77±13 years), with higher prevalence of hypertension (92.2% vs 85.9%) and diabetes (53.5% vs 28.6%). There was no difference between the two groups in terms of baseline eGFR, urine albumin, usage of ACEI (though there was more utilization of ARB 20.9% vs 15.7% in the obese group), non-potassium sparing diuretics or randomization to spironolactone. The group with obesity had a higher percentage of patients meeting NYHA class III and IV (39.3% vs 27.8%).

**Table 1:** Baseline Characteristics by BMI Group.

|  | **BMI<30** | **BMI≥30** | **P value** |
| --- | --- | --- | --- |
| N | 631 | 1134 |  |
| Age, years | 77 (13) | 69 (15) | <0.01 |
| Female | 300 (48.2) | 578 (51) | 0.27 |
| White Race | 512 (82.2) | 864(76.2) | <0.01 |
| Prior Smoking History | 305 (49) | 591 (52.1) | 0.24 |
| Hypertension | 534 (85.9) | 1046 (92.2) | <0.01 |
| Diabetes | 178 (28.6) | 607 (53.5) | <0.01 |
| ACEI | 178 (28.6) | 346 (30.5) | 0.38 |
| ARB | 98 (15.7) | 237 (20.9) | <0.01 |
| Diuretics | 337 (54.1) | 664 (58.6) | 0.07 |
| Randomization to Spironolactone | 304 (48.8) | 580 (51.1) | 0.36 |
| EF, % | 58 (12) | 59 (11) | 0.18 |
| Height, cm | 167.6 (15.3) | 167 (17.8) | 0.21 |
| Weight, kg | 72.6 (18.1) | 102.3 (28.9) | <0.01 |
| BMI, kg/m<sup>2</sup> | 26.3 (4.3) | 36.6 (8.3) | <0.01 |
| WC, cm | 96.5 (15.1) | 116.8 (20.3) | <0.01 |
| WHtR | 0.57 (0.08) | 0.70 (0.12) | <0.01 |
| sBP, mmHg | 122 (26) | 127 (23) | <0.01 |
| dBp, mmHg | 69 (16) | 70 (18) | <0.01 |
| NYHA class III or IV | 173 (27.8) | 445 (39.3) | <0.01 |
| Creatinine, mg/dL | 1.1 (0.5) | 1.1 (0.5) | 0.31 |
| Baseline eGFR, ml/min/1.73 m<sup>2</sup> | 60 (27) | 61 (29) | 0.58 |
| BUN, mg/dL | 22 (12) | 22 (13) | 0.67 |
| BNP, pg/mL | 498 (829) | 360 (535) | <0.01 |
| Urine albumin, mg/g* | 30 (101) | 27 (115) | 0.80 |
| Country |  |  |  |
| United States | 371 (59.6) | 775 (68.3) | <0.01 |
| Canada | 134 (21.5) | 192 (16.9) |  |
| Brazil | 74 (11.9) | 88 (7.8) |  |
| Argentina | 44 (7.1) | 79 (7.0) |  |

Values presented as either n (%), mean (standard deviation). The eGFR was calculated using the creatinine-based eGFR<sub>cr</sub> CKD-EPI 2021 equation
\*Among the subgroup with urine albumin available (n=1174)
BMI: body mass index; ACEI: angiotensin-converting enzyme inhibitor; ARB: angiotensin II receptor blocker; WC: waist circumference; WHtR: waist to height ratio; sBP: systolic blood pressure; dBp: diastolic blood pressure; NYHA: New York Heart Association; eGFR: estimated glomerular filtration rate; BNP: b-type natriuretic peptide; BUN: blood urea nitrogen; BNP: b-type natriuretic peptide

### Single eGFR Decline

A total of 686 (39.1%) patients reached the endpoint of single>30% eGFR decline. The mean frequency of eGFR measurements available during follow-up was 8±5 measurements. Kaplan-Meier curve analyses demonstrated significantly greater risk of >30% eGFR decline among individuals meeting obesity criteria based on BMI (P value = 0.029, Figure 1, panel A). A similar trend towards eGFR decline was observed in patients with obesity defined by WC criteria, although this did not reach statistical significance (P value = 0.089, Figure 1, panel B). No significant difference in the endpoint was detected among patients classified as obese by WHtR criteria (P value = 0.35, Figure 1, Panel C). There was no evidence of effect modification by randomization to spironolactone (interaction P values of 0.44 for BMI, 0.31 for WC and 0.95 for WHtR models).

**Figure 1.**
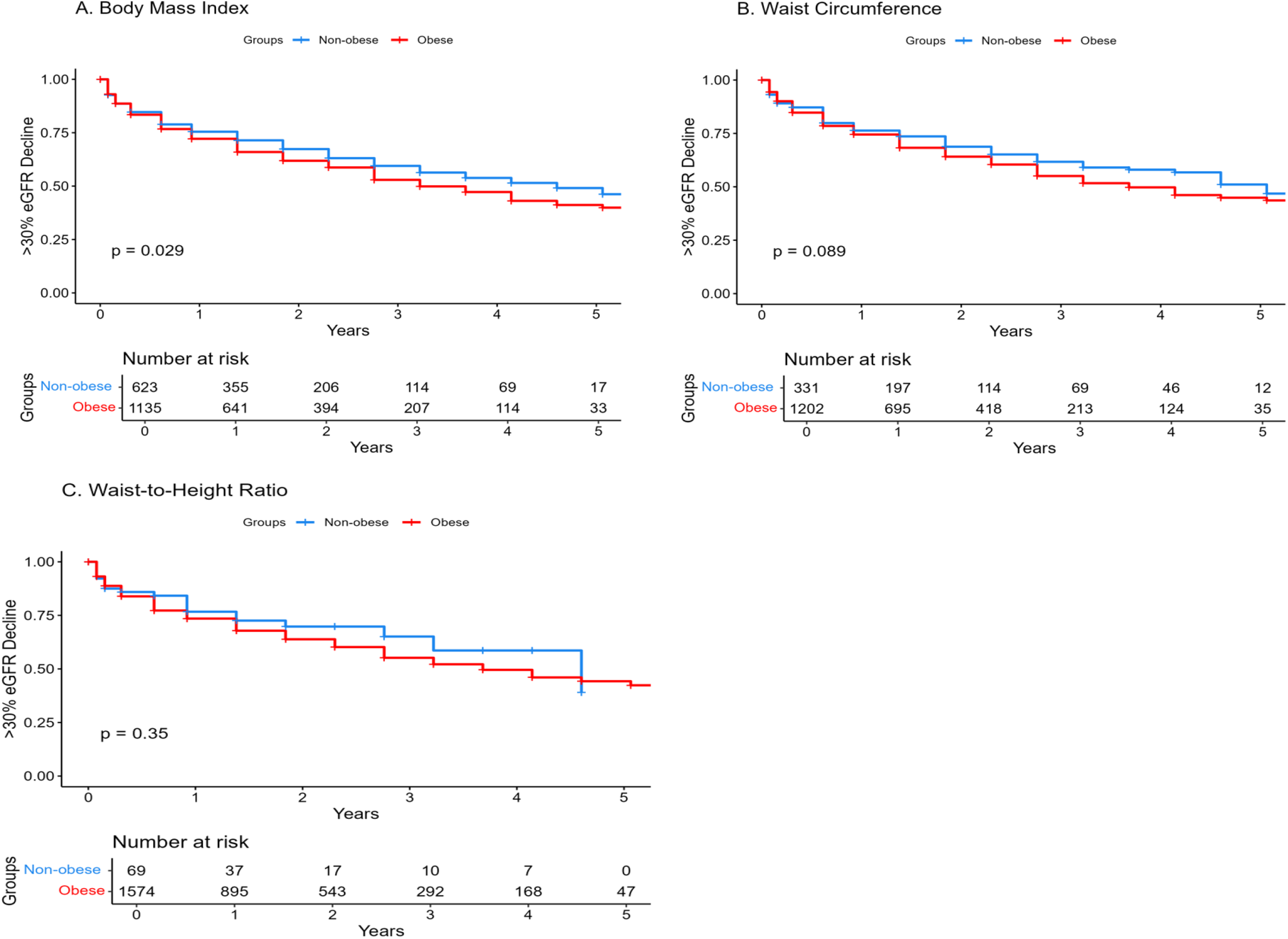
Kaplan-Meier curves for eGFR>30% decline for obese versus non-obese patients defined by BMI (Panel A), WC (Panel B) and WHtR (Panel C). The number at risk can be found below each separate plot. For BMI, the World Health Organization’s threshold of BMI ≥30 kg/m^2^ was used. For WC, previously used definitions of ≥102 cm in men and ≥88 cm in women were used.^6^ For WHtR, previously used definition of ≥0.5 was used. BMI: body mass index; WC: waist circumference; WHtR: waist to height ratio; eGFR: estimated glomerular filtration rate

When evaluated as continuous variables, BMI, WC and WHtR were each independently associated with the outcome after multivariable adjustment. Every SD higher in BMI was associated with a 10% higher risk of kidney function decline (HR 1.10 [1.01, 1.20]; Table 2). Similar associations were observed for WC (HR 1.10 [1.00, 1.20]; Table 3) and WHtR (HR 1.13 [1.04, 1.23]; Table 4).

**Table 2:** BMI: Univariable and Multivariable Analysis for eGFR 30% Decline.

|  | N of Event/N (%) | Unadjusted HR (CI) | Adjusted HR (CI)* | N of Event/N (%) | Unadjusted HR (CI) | Adjusted HR (CI)* |
| --- | --- | --- | --- | --- | --- | --- |
|  | Single 30% eGFR Decline |  |  | Persistent 30% eGFR Decline |  |  |
| Per unit increase in SD | 686/1756 (39.1) | 1.13 (1.05, 1.22) | 1.10 (1.01, 1.20) | 457/1767 (26) | 1.11 (1.01, 1.21) | 1.09 (0.99, 1.21) |
| Q1 $\leq 27.9 \text{ kg/m}^2$ | 150/440 (34.1) | Ref | Ref | 98/440 (22.3) | Ref | Ref |
| Q2 27.9-32.9 $\text{kg/m}^2$ | 173/439 (39.3) | 1.07 (0.86, 1.34) | 1.11 (0.89, 1.38) | 121/440 (27.5) | 1.11 (0.85, 1.45) | 1.17 (0.89, 1.53) |
| Q3 32.9-38.4 $\text{kg/m}^2$ | 171/439 (38.9) | 1.07 (0.86, 1.33) | 1.10 (0.88, 1.38) | 119/439 (27.1) | 1.14 (0.87, 1.48) | 1.20 (0.91, 1.59) |
| Q4 $>38.4 \text{ kg/m}^2$ | 193/438 (44.1) | 1.36 (1.10, 1.68) | 1.26 (1.00, 1.60) | 119/438 (27.2) | 1.20 (0.92, 1.56) | 1.11 (0.83, 1.50) |

\*Adjusted for age, sex, race, history of diabetes and hypertension, baseline eGFR, use of ACEI/ARB, diuretics and randomization spironolactone
BMI: body mass index; eGFR: estimated glomerular filtration rate; ACEI: angiotensin-converting enzyme inhibitor; ARB: angiotensin II receptor blocker;

**Table 3:** Waist Circumference: Univariable and Multivariable Analysis for eGFR 30% Decline.

|  | N of Event/N (%) | Unadjusted HR (CI) | Adjusted HR (CI)* | N of Event/N (%) | Unadjusted HR (CI) | Adjusted HR (CI)* |
| --- | --- | --- | --- | --- | --- | --- |
|  | Single 30% eGFR Decline |  |  | Persistent 30% eGFR Decline |  |  |
| Per unit increase in SD | 637/1641 (38.9) | 1.08 (1.00, 1.16) | 1.10 (1.00, 1.20) | 424/1642 (25.8) | 1.05 (0.96, 1.16) | 1.10 (0.98, 1.22) |
| Q1<sup>a</sup> | 148/411 (36) | Ref | Ref | 95/411 (23.1) | Ref | Ref |
| Q2 | 163/411 (39.7) | 1.14 (0.91, 1.42) | 1.12 (0.90, 1.41) | 115/412 (27.9) | 1.18 (0.90, 1.54) | 1.30 (0.99, 1.72) |
| Q3 | 148/413 (35.8) | 0.97 (0.77, 1.22) | 0.90 (0.71, 1.14) | 102/413 (24.7) | 1.00 (0.76, 1.32) | 1.06 (0.79, 1.41) |
| Q4 | 178/406 (43.8) | 1.37 (1.10, 1.71) | 1.35 (1.06, 1.71) | 112/406 (27.6) | 1.22 (0.93, 1.60) | 1.30 (0.97, 1.75) |

\*Adjusted for age, sex, race, history of diabetes and hypertension, baseline eGFR, use of ACEI/ARB, diuretics and randomization spironolactone
<sup>a</sup>Quartiles are sex specific with the following cutoff: Q1, women ≤93.9 cm; men ≤101 cm; Q2, women 93.9-106 cm; men 101-111.8 cm; Q3, women 106-116.8 cm; men 111.8-124.5 cm; Q4, women >116.8 cm; men >124.5 cm
WC: waist circumference; eGFR: estimated glomerular filtration rate; ACEI: angiotensin-converting enzyme inhibitor; ARB: angiotensin II receptor blocker;

**Table 4:** WHtR: Univariable and Multivariable Analysis for eGFR 30% Decline.

|  | N of Event/N (%) | Unadjusted HR (CI) | Adjusted HR (CI)* | N of Event/N (%) | Unadjusted HR (CI) | Adjusted HR (CI)* |
| --- | --- | --- | --- | --- | --- | --- |
|  | Single 30% eGFR Decline |  |  | Persistent 30% eGFR Decline |  |  |
| Per unit increase in SD | 637/1641 (38.9) | 1.15 (1.06, 1.24) | 1.13 (1.04, 1.23) | 424/1642 (25.8) | 1.13 (1.03, 1.25) | 1.12 (1.01, 1.24) |
| Q1 ≤0.59 | 142/411 (34.5) | Ref | Ref | 92/411 (22.4) | Ref | Ref |
| Q2 0.59-0.65 | 151/412 (36.6) | 1.01 (0.80, 1.27) | 0.98 (0.78, 1.23) | 101/412 (24.5) | 1.05 (0.79, 1.39) | 1.02 (0.76, 1.35) |
| Q3 0.65-0.73 | 167/409 (40.7) | 1.19 (0.95, 1.49) | 1.10 (0.88, 1.39) | 121/410 (29.5) | 1.37 (1.04, 1.79) | 1.29 (0.97, 1.70) |
| Q4 >0.73 | 177/409 (43.3) | 1.35 (1.08, 1.69) | 1.27 (1.00, 1.61) | 110/409 (26.9) | 1.25 (0.95, 1.66) | 1.18 (0.88, 1.58) |

\*Adjusted for age, sex, race, history of diabetes and hypertension, baseline eGFR, use of ACEI/ARB, diuretics and randomization to spironolactone
WHtR: waist to height ratio; eGFR: estimated glomerular filtration rate; ACEI: angiotensin-converting enzyme inhibitor; ARB: angiotensin II receptor blocker

When evaluated as quartiles, individuals in the highest quartiles of BMI, WC and WHtR exhibited significantly higher risk of eGFR decline. The most prominent association was seen with WC, where patients in the highest WC quartile had a 37% increased risk of single eGFR 30% decline in unadjusted models (HR 1.37 [1.10, 1.71]) with slight attenuation after covariate adjustment (HR 1.35, [1.06, 1.71]). Similar findings were noted for the highest quartiles of BMI (HR 1.26 [1.00, 1.60]) and WHtR (HR 1.27, [1.00, 1.61]).

### Persistent eGFR Decline

A total of 457 (26%) patients experienced persistent decline in eGFR>30%. Every SD increase in WHtR was associated with 12% increased risk of the outcome after adjustment for covariates (HR 1.12 [1.01, 1.24]). Similar trends were observed for BMI (HR 1.09 [0.99, 1.21]) and WC (HR 1.10 [0.98, 1.22]), although these association did not meet statistical significance.

Assessing evaluated as quartiles, most associations were attenuated. However, patients in the highest quartile of WC demonstrated a non-significant trend toward elevated risk (HR 1.30 [0.97, 1.75]) in the adjusted model.

### Subgroup Analyses

We performed subgroup analysis for men vs women, spironolactone vs placebo with regard to all three measures of obesity, including BMI, WC and WHtR.

In the gender subgroup analysis, significant associations between measures of obesity and single >30% eGFR decline were observed among women. Every SD increase in WC leads to 15% increased risk of the outcome (HR 1.15 [1.02, 1.31]), with similar findings seen for BMI and WHtR (Table 5). These association were not evident in men and were not observed for persistent eGFR decline in either group.

**Table 5:** Multivariable Cox Regression Analysis for Men and Women Subgroup for Single and Persistent eGFR 30% Decline.

|  | Men |  | Women |  |
| --- | --- | --- | --- | --- |
|  | Single eGFR Decline | Persistent eGFR Decline | Single eGFR Decline | Persistent eGFR Decline |
| **BMI per unit increase in SD** |  |  |  |  |
| N of Events | 312 | 203 | 374 | 254 |
| N | 883 | 883 | 882 | 882 |
| Event Rate (%) | 35.3 | 23.0 | 42.4 | 28.8 |
| Unadjusted HR | 1.16 (1.03, 1.31) | 1.14 (0.98, 1.32) | 1.10 (1.00, 1.20) | 1.07 (0.96, 1.20) |
| Adjusted HR* | 1.09 (0.94, 1.25) | 1.07 (0.90, 1.27) | 1.14 (1.02, 1.28) | 1.11 (0.98, 1.27) |
| **WC per unit increase in SD** |  |  |  |  |
| N of Events | 297 | 191 | 339 | 233 |
| N | 883 | 883 | 882 | 882 |
| Event Rate (%) | 33.6 | 21.6 | 38.4 | 26.4 |
| Unadjusted HR | 1.13 (1.01, 1.27) | 1.12 (0.96, 1.29) | 1.10 (0.98, 1.22) | 1.06 (0.93, 1.21) |
| Adjusted HR* | 1.10 (0.97, 1.25) | 1.07 (0.91, 1.27) | 1.15 (1.02, 1.31) | 1.12 (0.97, 1.30) |
| **WHtR per unit increase in SD** |  |  |  |  |
| N of Events | 297 | 191 | 339 | 233 |
| N | 883 | 883 | 882 | 882 |
| Event Rate (%) | 33.6 | 21.6 | 38.4 | 26.4 |
| Unadjusted HR | 1.19 (1.05, 1.34) | 1.19 (1.02, 1.38) | 1.10 (1.00, 1.22) | 1.07 (0.95, 1.21) |
| Adjusted HR* | 1.15 (1.01, 1.31) | 1.14 (0.97, 1.35) | 1.15 (1.03, 1.29) | 1.12 (0.98, 1.28) |

\*Adjusted for age, race, history of diabetes and hypertension, baseline eGFR, use of ACEI/ARB, diuretics and randomization to spironolactone
BMI: body mass index; WC: waist circumference; WHtR: waist to height ratio; eGFR: estimated glomerular filtration rate; ACEI: angiotensin-converting enzyme inhibitor; ARB: angiotensin II receptor blocker

In the spironolactone subgroup analysis, WHtR as a continuous variable was associated with elevated risk of single >30% eGFR decline in both placebo and spironolactone subgroup in the multivariable model (Table 6). Notably, WHtR also demonstrated a significant association with persistent >30% eGFR decline within the spironolactone group.

**Table 6:** Multivariate Cox Regression Analysis for Spironolactone and Placebo Subgroups for Single and Persistent eGFR 30% Decline.

|  | **Spironolactone** |  | **Placebo** |  |
| --- | --- | --- | --- | --- |
|  | Single eGFR Decline | Persistent eGFR Decline | Single eGFR Decline | Persistent eGFR Decline |
| **BMI per unit increase in SD** |  |  |  |  |
| N of Events | 419 | 295 | 267 | 162 |
| N | 886 | 886 | 879 | 879 |
| Event Rate (%) | 47.3 | 33.3 | 30.4 | 18.4 |
| Unadjusted HR | 1.10 (1.00, 1.21) | 1.08 (0.97, 1.21) | 1.17 (1.04, 1.32) | 1.16 (0.99, 1.35) |
| Adjusted HR* | 1.11 (0.99, 1.23) | 1.11 (0.97, 1.26) | 1.17 (1.02, 1.34) | 1.14 (0.95, 1.36) |
| **WC per unit increase in SD** |  |  |  |  |
| N of Events | 392 | 278 | 216 | 146 |
| N | 886 | 886 | 881 | 879 |
| Event Rate (%) | 44.2 | 31.4 | 24.5 | 16.6 |
| Unadjusted HR | 1.04 (0.94, 1.15) | 1.02 (0.91, 1.15) | 1.14 (1.01, 1.30) | 1.12 (0.95, 1.32) |
| Adjusted HR* | 1.06 (0.95, 1.18) | 1.04 (0.91, 1.18) | 1.13 (0.97, 1.30) | 1.09 (0.90, 1.32) |
| **WHtR per unit increase in SD** |  |  |  |  |
| N of Events | 392 | 278 | 244 | 146 |
| N | 886 | 886 | 879 | 879 |
| Event Rate (%) | 44.2 | 31.4 | 27.8 | 16.6 |
| Unadjusted HR | 1.15 (1.04, 1.27) | 1.12 (0.99, 1.26) | 1.17 (1.03, 1.33) | 1.17 (0.99, 1.37) |
| Adjusted HR* | 1.17 (1.05, 1.30) | 1.15 (1.01, 1.31) | 1.15 (1.00, 1.32) | 1.13 (0.95, 1.36) |

\*Adjusted for age, sex, race, history of diabetes and hypertension, baseline eGFR, use of ACEI/ARB, diuretics
BMI: body mass index; WC: waist circumference; WHtR: waist to height ratio; eGFR: estimated glomerular filtration rate; ACEI: angiotensin-converting enzyme inhibitor; ARB: angiotensin II receptor blocker

### Sensitivity Analyses

Exploratory analysis for single eGFR decline by >40% and persistent eGFR decline by >40% were performed (Supplementary Tables S3-S5). Most associations between measures of obesity as continuous variables and eGFR decline by >40% showed similar trends but were overall attenuated and no longer met statistical significance after adjustment. Among quartiles, the strongest association remained with obesity defined by WC. Patients in the highest quartile of WC had an increased risk of >40% eGFR decline (HR 1.40 [1.02, 1.92]) in the adjusted model. The number of patients meeting the strictest endpoint of persistent >40% eGFR decline decreased to 218 (13.3%). Trends remained similar albeit attenuated and without meeting statistical significance.

Results from competing risk analyses treating death prior to kidney outcomes as a competing event are shown in Supplementary Table S6. Associations between measures of obesity and eGFR decline were similar compared to results from the Cox regression models.

## Discussion

This study suggests that obesity and measures of abdominal adiposity are associated with increased risk of kidney function decline in patients with HFpEF, when defined as meeting a >30% decline in eGFR at one time during follow-up. The association attenuates but persists after adjustment for traditional risk factors including diabetes and hypertension as well as baseline eGFR. Furthermore, our findings suggest potential sex specific differences in the obesity-kidney relationship.

Patients with HFpEF have been described as having a higher prevalence of CKD than their heart failure with reduced ejection fraction counterparts, and presence of CKD is associated with numerous adverse outcomes including death, cardiovascular events and HF hospitalization.^11,12^ In light of the emerging cardio-kidney-metabolic syndrome paradigm of heart failure, obesity is materializing as having a key role in both pathophysiology and target for treatment in HFpEF.^13,14^

Population studies in the general non-HF patients have revealed that obesity can independently contribute to kidney function decline.^15^ A retrospective analysis of 320,252 adults followed over 15 years showed a three-fold increase in the relative risk of end stage kidney disease in class I obesity compared with persons with normal weight, and the association persists after adjustment for baseline blood pressure and diabetes.^16^ Similarly, another study observed that obese individuals with low or no metabolic risk factors had a higher risk of kidney function decline compared to normal weight metabolically healthy individuals.^17^ In contrast, limited studies exist on the effect of obesity on kidney function in the heart failure population. One study found that while obese patients with HFrEF had worse baseline renal function, higher BMI did not predict a more rapid decline in eGFR slope over a median of 2-years.^18^

In our study, obesity had a modestly increased risk of validated surrogate kidney outcomes, meeting statistical significance in the highest quartiles of BMI, WC and WHtR for single >30% eGFR decline. In most results, we also observed attenuation in the associations after adjustment. There are several possible explanations. The independent effect of obesity may be relatively small compared to coexistence of comorbidities such as diabetes and hypertension and the risk related to baseline eGFR. Secondly, the prevalent use of ACEI/ARB and spironolactone targets the RAAS, which is thought to be upregulated in obesity.^2^ Notably, randomization to spironolactone versus placebo in subgroup analysis did not show a significant difference on the impact of obesity on kidney function — consistent with FINEARTS-HF trial, where randomization to finerenone was associated with decreased risk of the cardiac composite outcome, but did not significantly alter kidney outcomes (HR 1.33 [0.94– 1.89]).^19^ While we also observed a higher number of kidney events in the spironolactone arm than the placebo arm, particularly for meeting the kidney event by a single eGFR value, this is likely due to spironolactone’s known acute hemodynamic decline in eGFR following initiation.^20^ We did not see any evidence that randomization to spironolactone had any clear modifying effect on the relationship between obesity and declines in eGFR.

The associations between measures of obesity and kidney function decline attenuated when using persistent kidney function decline as the endpoint. Such observations may be due to decreased power with a smaller number of events. Another interpretation could be that obese individuals with HFpEF may experience greater fluctuations in kidney function, rather than persistent decline. Prior studies have shown that obese patients with HFpEF demonstrate a greater rise in creatinine with decongestion compared with non-obese patients, as well as compared to those with HFrEF.^21,22^ These findings reveal the unique pathophysiology of HFpEF, and that obesity may amplify the underlying hemodynamic derangements. Furthermore, our hypothesis that obesity is a risk factor for greater hemodynamic-driven fluctuations in eGFR (that are not necessarily persistent) may help rectify the inconsistencies in the literature of obesity as an independent risk factor for kidney function endpoints.

We observed the most consistent associations between WC and declines in kidney function, defined both as a >30% and a >40% decline in eGFR at a single timepoint. There are several proposed mechanisms to explain the potential effect of abdominal obesity on kidney function. Locally, increased deposition of peri-nephritic fat can increase greater glomerular permeability, glomerular hypertrophy, leading to ultimate nephron loss.^23^ Individuals with perinephric fat had higher chance of CKD (OR 2.3, [1.28-4.14]) after adjustment for BMI and visceral obesity.^24^ Systemically, adipose tissue can induce production of leptin and adipokine, which directly causes RAAS activation.^2^ In addition to local and systemic effects on the heart, obesity induces an inflammatory state, which causes coronary endothelium to reduce production of nitric oxide and cGMP (cyclic guanosine monophosphate), and leads to myocardial hypertrophy, impaired relaxation and myocardial stiffness, further impairing ventricular dilation. ^4,25,26^ Markers of abdominal obesity have been shown to be stronger predictors of albuminuria, decreased kidney function, cardiovascular mortality, and perhaps the reason for why associations between WC and kidney function decline were more consistent in our study than those of BMI or WHtR. ^2,27^

We observed the effect of obesity on single eGFR >30% decline is consistent in women, but not in men. While this observation could be by chance due to the smaller numbers with subgroup analysis, the results align with a prior pooled population study that obesity confers a higher risk of kidney disease in women compared to men (RR =1.92 [1.78–2.07] vs 1.49 [1.36–1.63]; P=0.001).^28^ The literature on sex-specific risks remains mixed. For instance, a cohort study comprising 100,753 participants followed over 17 years reported increased risk of ESRD with higher BMI in men, but not in women.^29^ The underlying mechanisms remain unclear but may involve sex differences in body fat composition.^30^ Although men have more visceral fat, visceral adipose tissue in women have shown to have a stronger association with cardiometabolic outcomes.^31,32^ Our findings, which are only exploratory given the relatively small number, may reveal potential gender differences in obesity-kidney disease relationship which warrants further investigation.

Presidential advisory from the American Heart Association have called for enhanced obesity management in maintaining cardiovascular-kidney-metabolic health with lifestyle/behavioral, pharmacologic and surgical options.^13^ Our study may further suggest evidence that in HFpEF, obesity may increase susceptibility to fluctuations in eGFR, and the effect is significant in severely and morbidly obese individuals. With the advent of glucagon-like peptide-1 (GLP-1) agonists, post hoc analyses have shown decreased progression in eGFR decline among those randomized to GLP-1 agonists, at least among patients with diabetes.^33,34^ Additionally, in those without diabetes, semaglutide was superior to placebo in overweight and obese patients in reducing risk of cardiovascular death.^35^ Kidney outcomes could be a potentially interesting outcome to study in future trials of GLP-1 agonists in HFpEF.

### Strengths and Limitations

Our study has several limitations. The eGFR >30% and >40% decline are widely accepted surrogate endpoints in CKD literature, but their use has not yet been validated in HF populations. Urine albumin to creatinine ratio was excluded in the analysis due to missing data, but of the data available, there was not a prominent burden of albuminuria in TOPCAT (median 30 mg/g). Patients with eGFR<30 ml/min or serum creatinine >2.5 mg/dL were excluded from TOPCAT. Lastly, due to the observational nature of our study, there is a potential for confounding bias. Strengths include longitudinal measures of creatinine that were collected according to a pre-specified protocol. Evaluating the kidney endpoint by either a single measure or two persistent measures have also enabled us to distinguish acute declines from fluctuations from persistent declines. While there are no data available on body fat composition, we incorporated several measures of obesity, including WC, which can be more specific to central obesity than BMI. Consideration of death prior to kidney outcome was taken into our sensitivity analyses, without large differences in results.

## Conclusion

Among ambulatory patients with HFpEF, there is a high burden of clinically significant declines in kidney function. Obesity was associated with higher risk of declines in kidney function, in particular acute eGFR declines and less so with persistent eGFR decline. With the promise of interventions such as GLP-1 agonists, the role of obesity as a risk factor for declines in kidney function may represent another potential target for intervention among patients with HFpEF.

## Data Availability

All data and materials have been made publicly available through the NHLBI's Biolincc data repository.

## Abbreviations

HFpEF: heart failure with preserved ejection fraction
LVEF: left ventricle ejection fraction
CKD: chronic kidney disease
eGFR: estimated glomerular filtration rate
NYHA: New York Heart Association
BMI: body mass index
WC: waist circumference
WHtR: waist to height ratio
NT-proBNP: N-Terminal Pro-B-Type Natriuretic Peptide

## Acknowledgements

The manuscript was prepared using data set provided by the NHLBI and does not necessarily reflect the opinions or views of TOPCAT or the NHLBI.

